# Excess mortality during COVID-19 in five European countries and a critique of mortality data analysis

**DOI:** 10.1101/2020.04.28.20083147

**Authors:** J. Félix-Cardoso, H. Vasconcelos, P. Pereira Rodrigues, R. Cruz-Correia

## Abstract

**INTRODUCTION:** The COVID-19 pandemic is an ongoing event disrupting lives, health systems, and economies worldwide. Clear data about the pandemic’s impact is lacking, namely regarding mortality. This work aims to study the impact of COVID-19 through the analysis of all-cause mortality data made available by different European countries, and to critique their mortality surveillance data.

**METHODS:** European countries that had publicly available data about the number of deaths per day/week were selected (England and Wales, France, Italy, Netherlands and Portugal). Two different methods were selected to estimate the excess mortality due to COVID19: (DEV) deviation from the expected value from homologue periods, and (RSTS) remainder after seasonal time series decomposition. We estimate total, age- and gender-specific excess mortality. Furthermore, we compare different policy responses to COVID-19.

**RESULTS:** Excess mortality was found in all 5 countries, ranging from 10.6% in Portugal (DEV) to 98.5% in Italy (DEV). Furthermore, excess mortality is higher than COVID-attributed deaths in all 5 countries.

**DISCUSSION:** The impact of COVID-19 on mortality appears to be larger than officially attributed deaths, in varying degrees in different countries. Comparisons between countries would be useful, but large disparities in mortality surveillance data could not be overcome. Unreliable data, and even a lack of cause-specific mortality data undermine the understanding of the impact of policy choices on both direct and indirect deaths during COVID-19. European countries should invest more on mortality surveillance systems to improve the publicly available data.

## 1. Introduction

2020 started with news about a strange pneumonia virus in Wuhan, China. Since then, it has been declared a Pandemic by the World Health Organization (WHO), and it has caused, as of April 19th, 2 281 714 confirmed cases and 159 511 deaths of COVID-19, of which 1 018 221 cases and 159 511 deaths in Europe [1].

COVID-19 is a clinical entity caused by a novel coronavirus, SARS-CoV-2. Its epidemiology is still uncertain. Person-to-person spread of SARS-CoV-2 is thought to occur mainly via respiratory droplets [2]. Mean incubation period ranges from 4 to 6 days and the mean serial interval ranges from 4 to 8 days [3]. There is probably an early peak of infectiousness, with probable presymptomatic transmission for some cases [4]. Median time from onset of symptoms to intensive care unit (ICU) admission is around 10 days [5]. WHO reported that the time between symptom onset and death ranged from about 2 weeks to 8 weeks [6]. Case Fatality Rate is estimated to range from 0.87 to 9.36 (updated 21^st^ April) [7].

Its novelty and characteristics make this virus’ impact hard to measure, and a lack of comparable and reliable data is not helping. The number of officially confirmed cases is highly dependent on testing policy and capacity. Some countries only test patients in need of hospitalization. Others have policies that recommend testing everyone that presents with a set of symptoms, regardless of their need of hospital care. However, to our knowledge, no country has been able to test every patient with suggestive clinical presentation, due to a global shortage of tests. Thus, the number of infected individuals will inevitably be larger than the number of confirmed cases of COVID-19.

Deaths attributed to COVID-19 are also difficult to measure accurately. Problems with testing affect not only confirmed cases but also attributed deaths. Besides, testing is not enough to determine the cause of death, as some patients may die while infected with SARS-CoV-2, but not due to it. Due to constraints health systems are facing across the world, it is likely that a precise attribution of cause of death is not possible. Additionally, some patients die without there having been a suspicion of COVID-19.

We postulate that mortality surveillance can be of help in the everyday management of health systems, but especially during health crisis such as infectious disease outbreaks where an estimate of observed excess mortality could be informative. Compared to the ability to identify, test and attribute deaths to a novel pathogen, measuring mortality is relatively simple. Furthermore, contrary to novel pathogens, which may have specificities that require the collection of unusual data, it benefits from the possibility of having IT systems in place in advance.

The EuroMOMO network monitors weekly all-cause age-specific excess mortality in countries in Europe through a standardised approach. At the moment, data is available for up to the 12^th^ of April (week 15). In week 11, Italy had high excess mortality, and it has had very high excess since then. Portugal had “above expected” mortality in weeks 13 and 14. The Netherlands faced high mortality in week 12 and very high ever since, and England as well. For Wales, week 13 had high mortality, and week 14 very high. Lastly, France had high mortality in week 12, very high in weeks 13 and 14, but no excess this last week. Beyond this qualitative analysis, we found four studies of excess mortality during COVID-19. Switzerland has a mortality surveillance system in place with data from the Federal Statistical Office [8] that estimates 892 excess deaths for those above-65 up to April 12th. Instituto Carlos III in Spain found a 56,5% all-cause mortality excess from March 17^th^ - April 7^th^ (13 954 excess deaths), mostly in individuals older than 75 and males [9]. Researchers familiar with the Spanish mortality surveillance system mentioned some concerns about data reliability for the last few days, so it’s likely that this may be revised upwards [10]. Cancelli and Foresti found increases in overall mortality in several Italian regions ranging from 4 to 10 times the amount of COVID-19 reported deaths [11]. They conclude that deaths attributed to COVID-19 are underestimated, and end their paper calling for more, better data on the pandemic. Galeotti et al. use a self-titled “rule of thumb calculation” pointing to a 1:1 to 1:3 COVID-19 to unexplained excess death count, a number not meant to be interpreted as an epidemiological model, but only as an insight into a potentially disregarded information gap [12]. These last two studies have strong limitations, most of all regarding the estimation of excess deaths, but it’s reasonable to assume that their conclusions stand.

Our aim is to quantitatively characterize all-cause mortality in selected European countries in an effort to better measure the impact of COVID-19, and to critique mortality surveillance data. We also confront mortality data with policy decisions.

## 2. Methods

We estimated excess mortality during COVID-19 in 5 European countries, using two different methods and comparing it to COVID-19 attributed deaths, and analysed policies undertaken by each country.

**DATA SOURCES** We searched for daily or weekly data on all-cause mortality for most European countries, particularly those participating in EuroMOMO. We could find suitable data for Portugal, Italy, France, England and Wales, and the Netherlands. We define suitable data as data available for a period matching the start of the COVID-19 outbreaks in Europe, publicly available, timely updated, and with daily or weekly resolution. Datasets are described in Table 1.

**Table 1:**
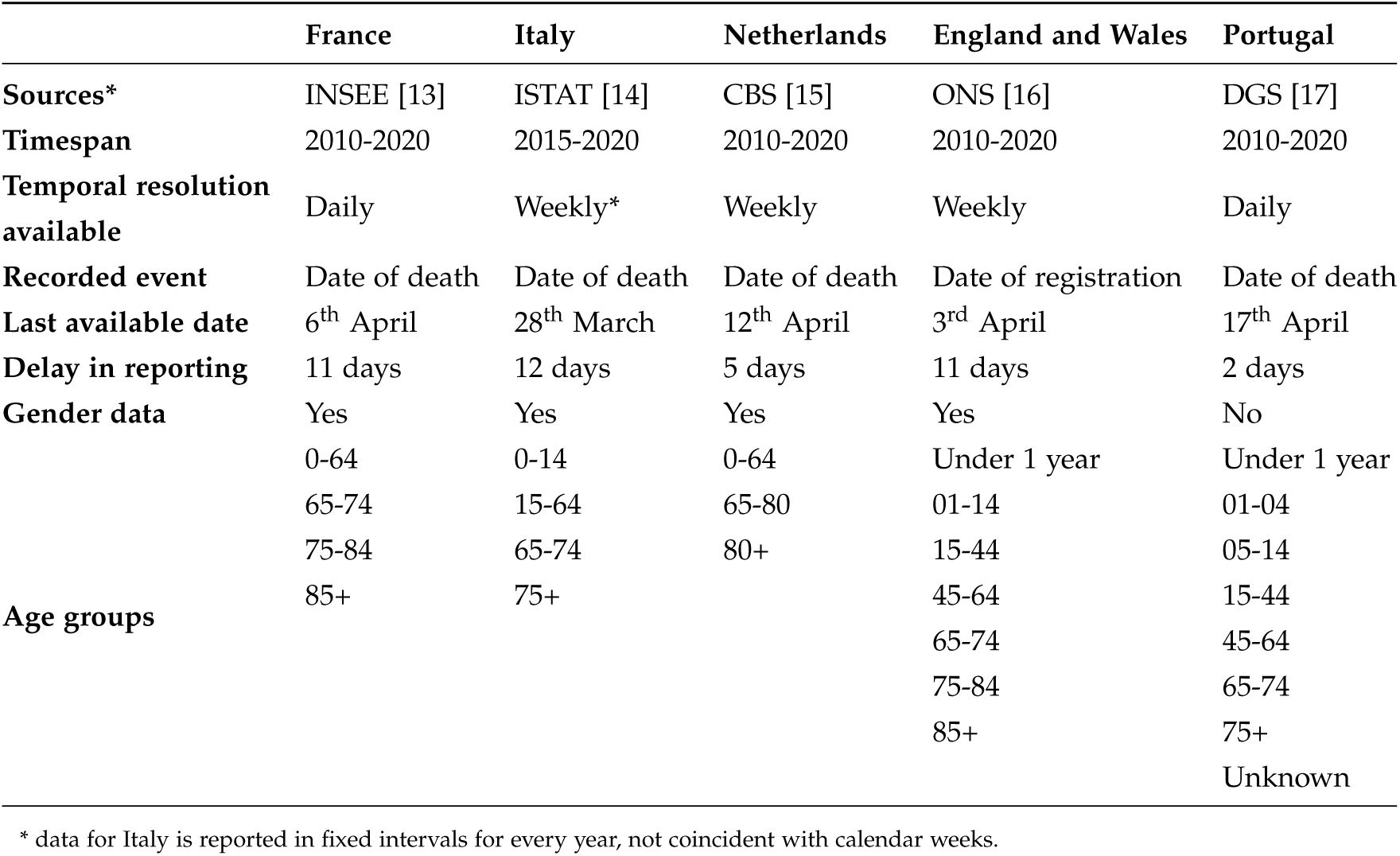
Data sources and characteristics.

We checked every dataset for internal consistency, comparing age and gender-specific totals and checking for strange values comparing different years/datasets available.

We collected data on COVID deaths from Johns Hopkins University CSSE Repository [18], except for England and Wales, where we used data from ONS [19] to use date of registration, as use for all-cause mortality.

**DATA HARMONIZATION** Datasets selected for this work are diverse and required some harmonization work. We can divide them according to several characteristics. Portugal and France have daily data, which we have decided to adapt into weekly data to have a common data unit and smoothen daily variations that may disturb the analysis. Italy, the Netherlands, and England and Wales already have weekly data. All countries report their weekly mortality differently, with the Netherlands using Monday as the first day of their week, England and Wales using Saturday, and Italy using a first, 11-day week, and 7-day weeks for the rest, regardless of the day of the week (and with one 8-day week for bissextile years). We used England and Wales as a template to adapt Portuguese and French data. Regarding age-groups, all countries report it differently, with under and over-65 as the only age group with comparable data. Thus, we created a dataset with age-specific mortality for under- and over-65 for all 5 countries. We have corrected Italian data for the last week of February, since they have 8-day weeks every bissextile year. We have considered only 7/8 of deaths reported for that week, both for 2016 and 2020. Stratified data from the Netherlands presented an artifact every year-end, which resulted in some years ending with 53 weeks (last one incomplete). For these, data from week 53 was aggregated with week 1 (also incomplete) from the following year. Portugal has no data on gender mortality. Our datasets are accessible on GitHub [20].

**DATA ANALYSIS** We calculated excess mortality based on two different statistical methods, which were applied to total all-cause mortality and to four subtotals: under-65, male and female, and over-65, male and female. We started our analysis on the day of the first COVID-19 death in each country, naming the first full week after that “week 1”. We calculated the amount of excess deaths that could be explained by COVID-19, and we also looked at week-to-week trends in total mortality for each country, using “week 0” as a baseline to chart a comparative evolution of weekly mortality in each country. Deviation from the expected value from homologue periods (DEV) Mortality is relatively stable across the years for all analyzed datasets. Thus, we calculate the expected mortality using the homologue mean for previous years and adding a standard deviation to account for normal variation. This may lead to an underestimation of excess deaths, as it sets a high threshold. Other authors use a higher threshold (3 z-scores for the EuroMOMO algorithm, for instance) but usually discount previous periods of excess mortality.

For this method, we present the results as the number of excess deaths for the timespan beginning with the first full week since the country’s first death, and consider only weeks with excess deaths for computing excess deaths as a percentage of expected deaths.

Remainder after seasonal time series decomposition (RSTS) The second method we used for estimating excess mortality considers the series of weekly number of deaths explicitly as a time series and applies a seasonal decomposition [21] to adjust observed mortality for seasonality and trend, exposing an irregular component (remainder) that should be an indication of the excess mortality in that period. It works by using LOESS [22] polynomial smoothing iteratively on the seasonal sub-series, after which the remainder is smoothed to find the trend. The remainder component is the residual from the seasonal plus trend fitted using weighted least squares. Remainders were found using a yearly seasonality and a 10-years long trend, using two models for each country: one where only the trend is explicitly included in the regression, and another where seasonality was also explicitly included in the regression. R was used to perform the data and statistical work, including the STL function of package ‘stats’ (R software, version 3.6.2, R Foundation for Statistical Computing). Results for RSTS are presented as an interval (defined by the result of both models), and the entire analysed time period is considered for reporting excesses in mortality. Analysis for different age and gender strata was also performed (if specific data were available). This method was not applied to Italy, since its dataset is not a year-long dataset but contains data just for the January-April period.

**POLICY ANALYSIS** We use the Oxford Covid-19 Government Response Tracker [23] and its Stringency Index to extract a date for main policy decisions in all five countries, and compare it to the timeline for all-cause mortality. Like elsewhere in this work, we use the 1^st^ COVID-19 death as a reference. We chose three policies as the most relevant: school closure, cancelling of public events, and domestic travel restrictions.

## 3. Results

**Data quality** Data quality is particularly contrasting among data sources. Internal coherence is lacking in some datasets. For Portugal, on 6^th^ March 2020 the sum of the district specific mortality for all districts does not equal the national total for the day, the only such mistake we were able to find. French data included deaths with no attributed date of death, which were naturally excluded from the total. These accounted for a total of 204 out of 5,988,702 entries (0.0034%). Italy also reports very incomplete data: only 1450 of the 7904 Italian counties. England and Wales have missing data on subtotal deaths (unknown age or gender). We also found poor data structure for some countries. Italy groups the first 11 days of the year together as a “week”, and uses the same calendar days for every year, even for bissextile years. Dutch datasets presented with ambiguous information regarding weeks at the end of a year which had less than 7 days: some years would draw on the truncated weeks for the previous year and solely update the rest of the days as their first week death count, and some others would present a total for a 7 day period. We did our best to mitigate the impact of these data quality issues with corrections as light as possible.

**Mortality trends** As seen in Figure 1, mortality does not evolve the same way since week 1 of COVID-19 in all countries. For most countries, week 1 is still close to normal, but week 2 starts showing increases in all-cause mortality. Largest week-on-week (WoW) increases happen on week 2 in Portugal and Italy, week 3 in the Netherlands, and week 4 in England and Wales. In France, the largest increase in mortality WoW happens on week 7.

**Figure 1:**
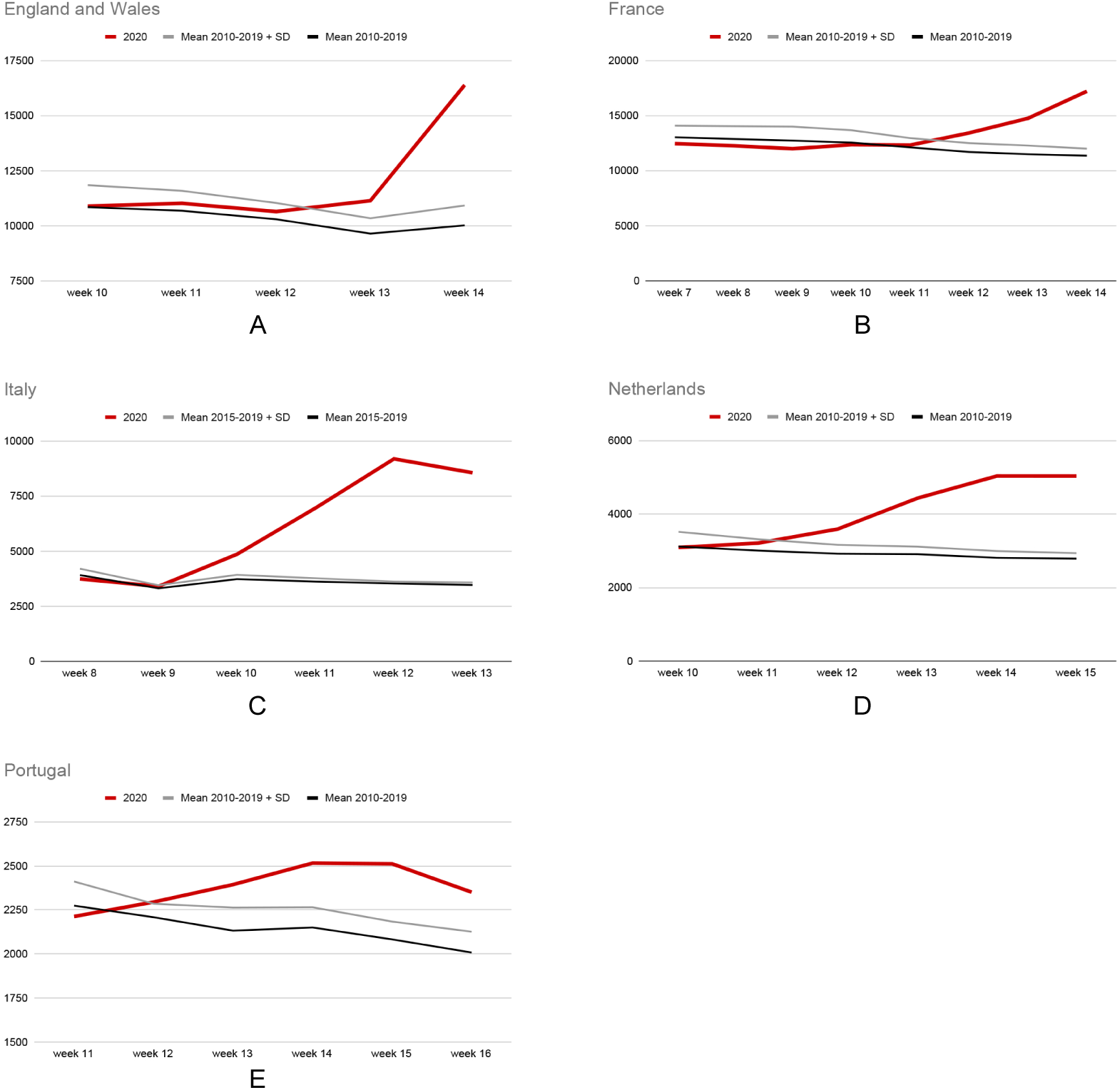
All-cause mortality data for selected countries, mean and 2020.

Table 2 presents data on excess deaths with breakdown by age and gender when possible, alongside the amount of deaths attributed to COVID-19, according to the DEV method. We found excess deaths in all studied countries. Italy stands out from the rest for its large excess mortality, almost doubling the upper limit of expected deaths. The percentage of excess deaths explained by COVID-19 in weeks of increased mortality varies substantially, from almost all excess deaths (England and Wales, 92.9%) to less than half (Netherlands, 46.1%). Age and gender differences were found, with excess mortality predominantly in elderly males, and with Portugal and France showing no excess mortality in under-65 populations.

**Table 2:**
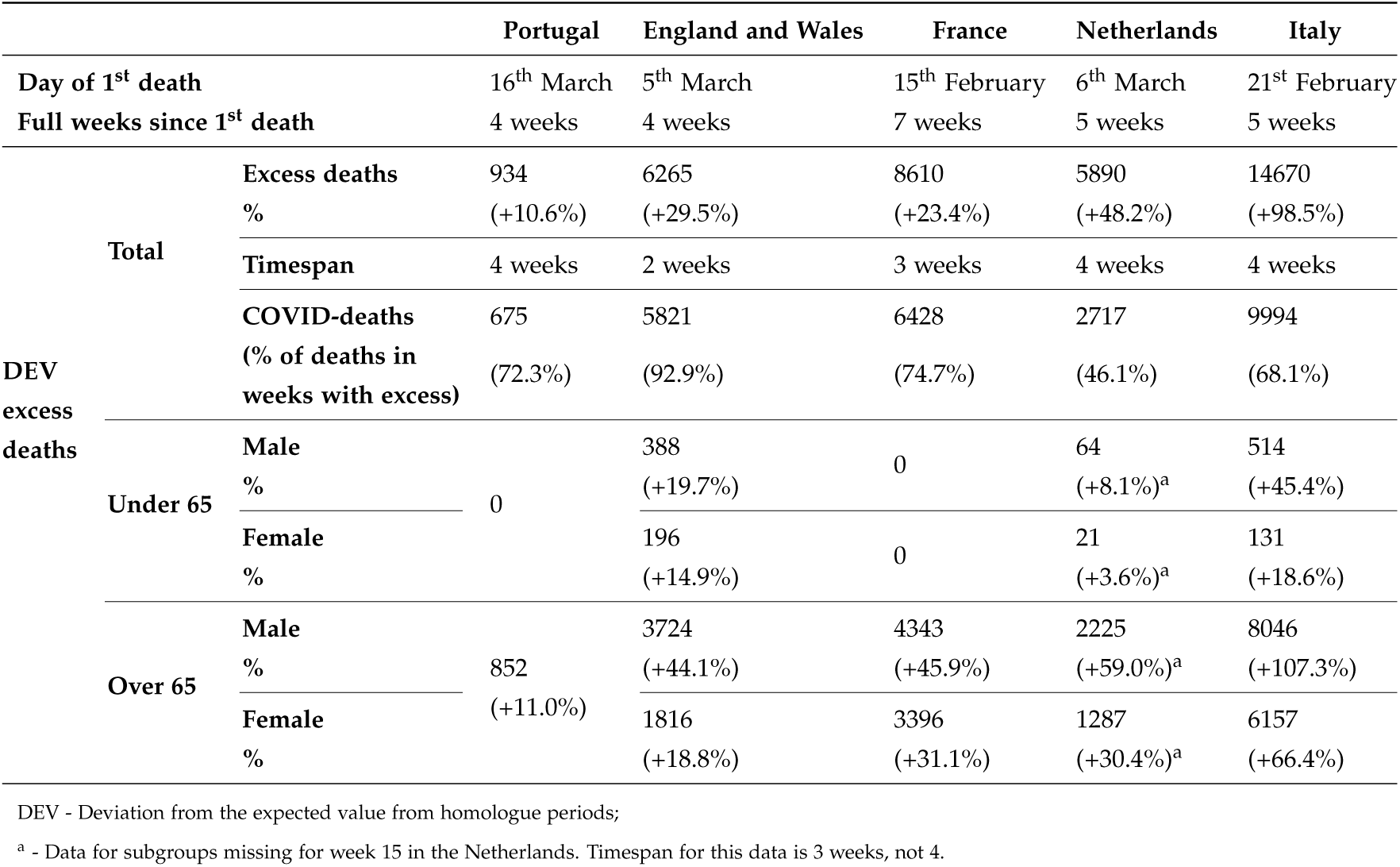
Total, age- and gender-specific excess mortality results with DEV method, selected countries, February-April 2020.

Table 3 presents data on excess deaths with breakdown by age and gender according to the RSTS method. Results are similar to the DEV method. Excess mortality can be observed in all 4 countries, ranging from single digit increases for France and Portugal to double digit increases in England and Wales and the Netherlands. As with the DEV method, we find excess mortality happens mostly in males over 65. Both France and Portugal present no excess deaths for under-65.

**Table 3:**
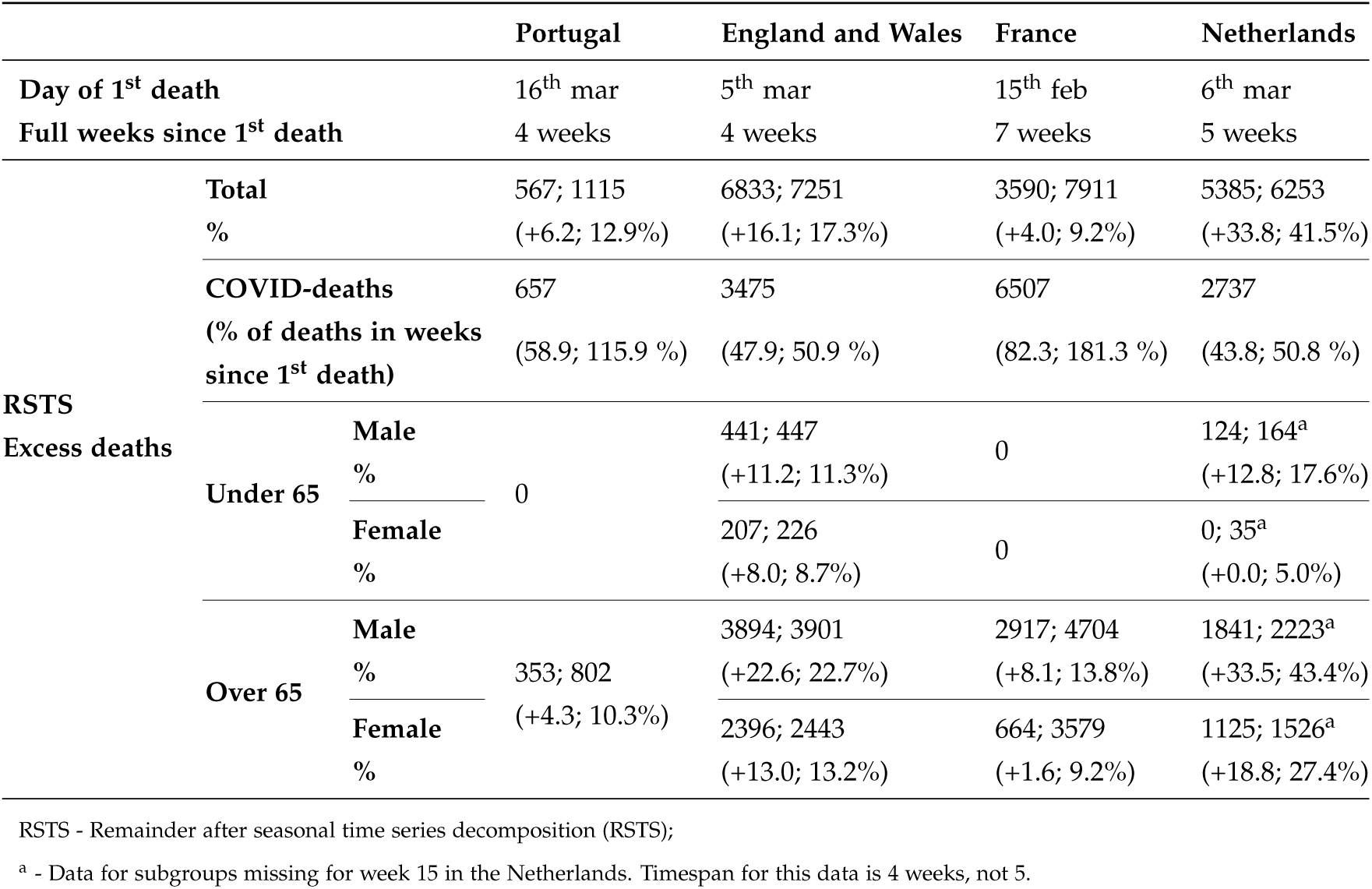
Total, age- and gender-specific excess mortality results with RSTS method, selected countries, February-April 2020.

Looking at Figure 2, we can divide countries in two groups: those whose mortality is still growing - France and England and Wales - and those who are currently experiencing a plateau or a decrease - Netherlands and Portugal, and Italy, respectively. Growth happens earlier in the Netherlands, Italy and Portugal, and later in England and Wales. Italy is a clear outlier, with 2 weeks of extreme excess mortality, more than doubling the baseline of week 0. The country who acted earlier - Portugal - presents the best results with week 4 showing a downward trend towards values similar to baseline mortality, and a smaller amount of excess deaths, both in absolute and relative terms.

**Figure 2:**
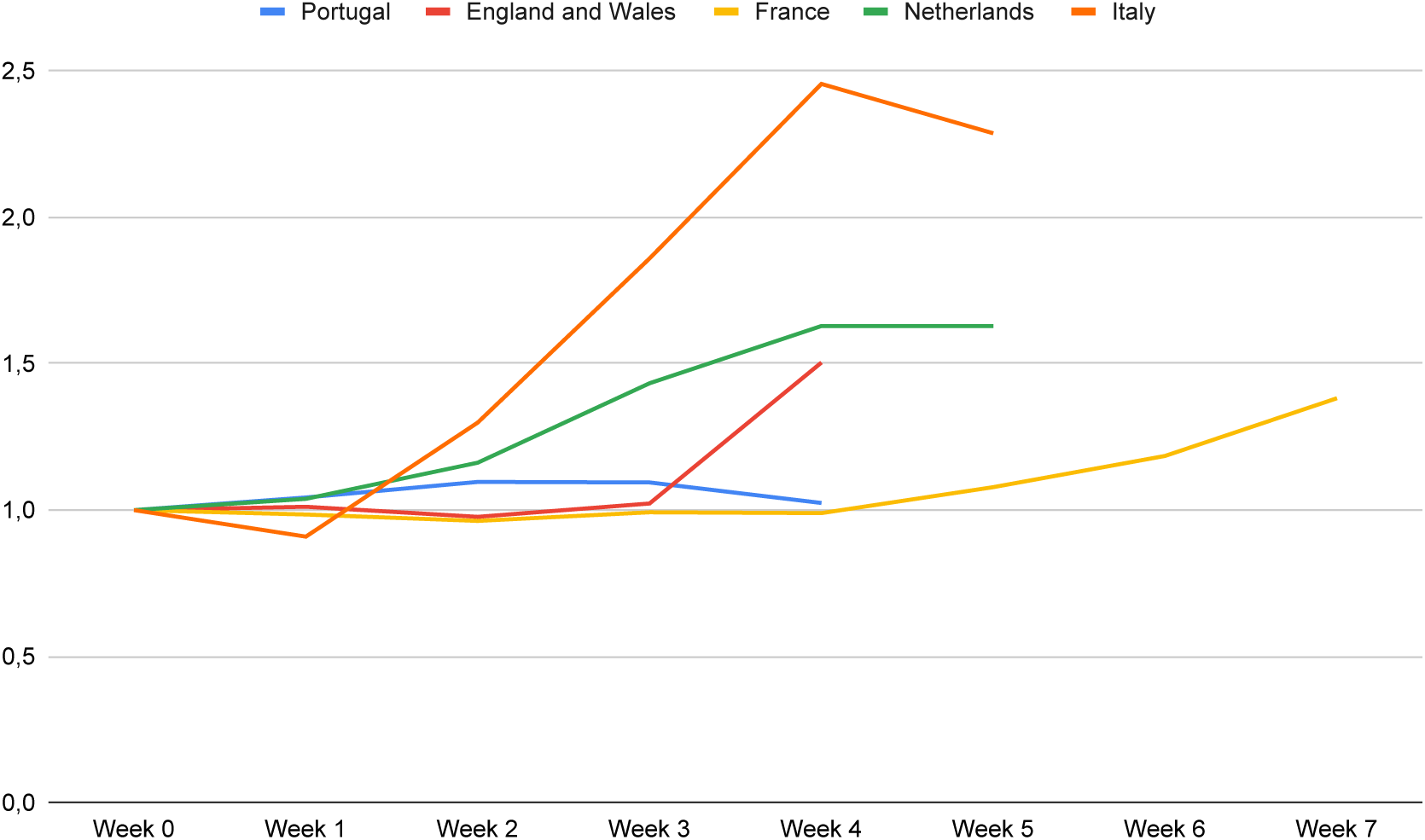
All-cause mortality trends, week 0 as baseline for all selected countries. Week 0 is the week before the 1^st^ full week after the first COVID-19 death in each country

## 4. Discussion

Our main aim was to study all-cause mortality data across different European countries. Specifically, we aimed to characterize mortality during COVID-19, including excess mortality, to compare different policy options and possible impacts on mortality, and to describe and critique mortality surveillance data.

All 5 studied countries present excess mortality during COVID-19, with large differences between the one who presents only a slight increase, Portugal, and those with large increases, such as Italy and England and Wales. Plus, excess mortality is larger than the amount of COVID-19 attributed deaths in all 5 countries, raising questions about the causes of excess mortality and about the reliability of cause of death attribution for such a novel and undertested disease. For some countries, it is particularly hard to understand how much of the excess mortality can be directly attributed to COVID-19, since deaths occur in large amounts outside hospitals. COVID-19 deaths happening in care homes range from 42% in Belgium to 57% in Spain [24]. Besides untested COVID-19-caused deaths, which are not quantified, other deaths may be happening due to the disruption of societies and health systems. Additionally, lower mortality rates during January and February, mostly due to a warm winter season in Europe and a light influenza season, may help explain some of this excess mortality, in an effect commonly called harvesting.

As would be expected considering data from COVID-19 deaths, the largest increase in mortality is felt in the elderly. Men over 65 are the most affected population group, with around twice the increase in deaths compared to that of women over 65. This may reflect a higher degree of frailty in elderly men, compared to elderly women, when exposed to the direct and indirect effects of the pandemic.

Indirect effects of the COVID-19 pandemic are felt due to, among others causes, a shift in healthcare resources and fund allocation towards pandemic containment. Acute care patients may also be less likely to go to an A&E service, fearing a higher risk of contagion. This would be confirmed by data from A&E. Poor strata of society may see communitarian help they rely on severely hampered. Conversely, some other changes due to COVID-19 reaction by the authorities may result in saved lives: traffic reduction decreases the chances of fatal road casualties; social distancing measures will lower the number of risky outdoors activities; a reduction of working hours leads to a decrease in chances of job accidents; and increased hand and respiratory hygiene decreases the rate of other infections, including influenza and other respiratory tract infections.

We compare our method with EuroMOMO for previous periods of the year to check our threshold against theirs, as EuroMOMO is a well-calibrated international tool [25]. EuroMOMO alerts for excess mortality in 2020 in Portugal (above normal (AN) in week 1 and 2, and high excess (HE) mortality in week 3), in England and Wales (AN in week 1), in the Netherlands (AN in week 2) and in Italy (AN in week 3). Our DEV method returns no excess mortality for any of these weeks. Our RSTS method reports increased mortality for week 3 in England and Wales, but no further periods of excess mortality. We find this to be sufficiently coherent to state that we have a higher threshold than EuroMOMO and are not overestimating excess mortality.

Countries who acted earlier relative to their first COVID deaths seem to have had better results for overall mortality. This is the case for Portugal, which has seen a smaller increase in overall mortality and a faster return to baseline mortality. However, poor data quality makes it difficult to propose any relationship between policy and mortality. Data for England and Wales must be interpreted carefully, since it does not represent date of death, but date of registration, which can account for such a delayed increase in excess mortality. France provides data with low reliability, as can be confirmed through the extensive delays in the reporting of deaths - the file for March, available in April, added deaths to January and February, thus making it possible that we’ll have to wait until June to truly quantify the mortality in France during the month of March. These two countries are the ones who present a delayed growth in mortality, which may be an artefact due to poor data quality.

### i. Relevance of this work

COVID-19 cases and mortality have been heavily scrutinized by public opinion. There has been great interest in this topic in the media, with some articles mentioning the unreliability of data thus far [26]. Scientists in some European countries have asked for access to better data in order to help fight the pandemic. This includes efforts to improve open data initiatives [27], [28], but also collaboration between government and academia in topics in which open data is not possible (detailed clinical records, for instance).

No country has publicly available mortality surveillance data accurate and detailed enough to answer important questions during the ongoing pandemic. The one who comes closest is Portugal, with a shorter delay, and more coherent and more refined data, but without detailed gender-specific or regional data and lacking causes of death. Monitoring excess mortality and its causes would also allow for a better balance between avoiding damage from the pandemic and damage caused by the measures taken to fight the pandemic. Some policymakers worry about the impact of social distancing and lockdown measures, and having up-to-date mortality data would be a useful tool to achieve the difficult balance between fighting an outbreak and avoiding larger than needed negative impacts for the population.

Despite the existence of regional data in most surveyed countries, it faces increased data problems compared to nationwide data. Italy has timely available data for less than half its regions; Portugal is only updating its “Districts”, which do not geographically match the health regional administrations that are responsible for COVID-19 data reporting. England and Wales, the Netherlands and France have detailed regional data, but it suffers from the same constraints as country data.

Appropriate EU-level guidance on mortality surveillance and reporting would solve many of the problems we identify. Recently, the EU Commission has adopted a toolbox for coordination on mobile contact tracing apps [29]. Despite this being an important step, we find it more useful to have coordinated COVID-19 case and death reporting, as well as allcause mortality data. Without accurate, pan-European, data, it is difficult to compare different policy strategies and prioritize resources for most-affected regions. This is particularly important considering the wide variety of policy responses during this crisis, which will be difficult to study without comparable, reliable, transparent data.

This study has several limitations related to the quality of data or methods that are important to mention. As previously stated, data has many problems and this greatly limits its interpretations. For this reason, limitations cannot be overcome, and thus we chose to be conservative in estimating excess mortality, as one can confirm when comparing our results to other results, such as those mentioned in this paper.

Regarding future work, we aim to include a regional analysis of each country and to add other countries. Data on causes of death would also help us understand collateral damages from the pandemic. Lastly, collecting data on hospital usage, especially A&E services, would provide good clues to whether excess deaths from non-COVID-19 causes could be avoided by strengthening other health services.

### ii. Main findings and recommendations

An excess of mortality was found in all studied countries in the period after the first COVID-19 attributed death, beyond those deaths directly confirmed as COVID-19. However, mortality surveillance systems in the five studied countries presented several data quality issues that hindered a more in-depth analysis, relevant both for pandemic and normal contexts. Therefore, as members of the international community of researchers that seek to work on European COVID-19 issues, the following recommendations are highly suggested:

1. Very few European countries have up-todate, publicly available, mortality data. Each European country should have a public mortality surveillance site (Portuguese site EVM is a good template)
2. Some countries take weeks to publish their mortality data, making it difficult to follow the evolution and analyse data in time to help decisions. The delay to make public data available should be the minimum possible (< 5 days)
3. Each country uses a different rule to group data into time periods (deaths per day; per week, starting in different weekdays, and with weeks having more than seven days in one country). Share data on daily number of deaths (not just grouped in weeks)
4. Relevant hypotheses are very difficult to test due to a lack of data, namely regarding causes of excess mortality. Add causes of death, using an international terminology standard, to the reports
5. Gross errors can be found in available data Improve internal automatic verification methods to detect and correct data problems before publishing data.

**Table 4:**
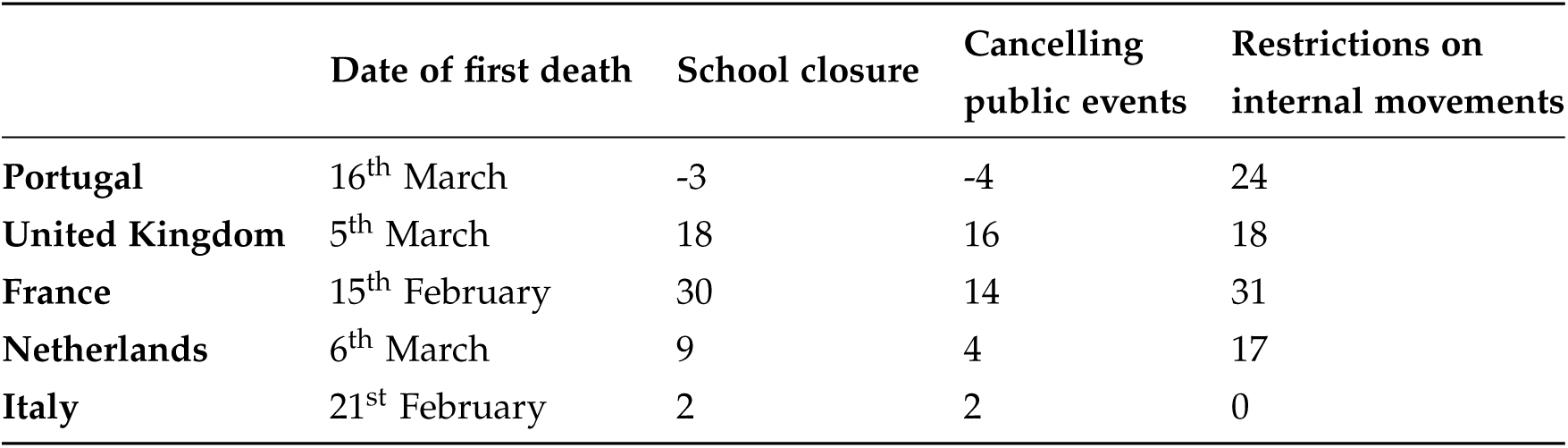
Policies enacted by countries relative to time of first death, days.

|  | Date of first death | School closure | Cancelling public events | Restrictions on internal movements |
| --- | --- | --- | --- | --- |
| **Portugal** | 16<sup>th</sup> March | -3 | -4 | 24 |
| **United Kingdom** | 5<sup>th</sup> March | 18 | 16 | 18 |
| **France** | 15<sup>th</sup> February | 30 | 14 | 31 |
| **Netherlands** | 6<sup>th</sup> March | 9 | 4 | 17 |
| **Italy** | 21<sup>st</sup> February | 2 | 2 | 0 |

## Data Availability

All data is available in a GitHub repository

https://github.com/henriquetguedes/ExcessMortalityCOVID

